# Elevated DNA Methylation Gestational Age is Associated with the Risk of Later Bipolar Disorder and Anorexia Nervosa in Twins

**DOI:** 10.1101/2020.07.16.20155010

**Authors:** Christine Søholm Hansen, Anna Starnawska, Alexander Werner Drong, Shantel Marie Weinsheimer, Marie Bækvad-Hansen, Dorte Helenius, Marianne Giørtz Pedersen, Carsten Bøcker Pedersen, Preben Bo Mortensen, Michael Christiansen, David Michael Hougaard, Cecilia Lindgren, Thomas Mears Werge, Jonas Bybjerg-Grauholm, Alfonso Buil Demur

**Affiliations:** The Lundbeck Foundation Initiative for Integrative Psychiatric Research, iPSYCH, Denmark; Department for Congenital Disorders, Statens Serum Institut, Copenhagen, Denmark; Institute of Biological Psychiatry, MHC Sct. Hans, Mental Health Services Copenhagen, Roskilde, Denmark; Department of Biomedicine - Health, Aarhus University, Aarhus, Denmark; Big Data Institute, University of Oxford, Oxford, United Kingdom; National Centre for Register-based Research, Department of Economics and Business Economics, Aarhus University, Aarhus, Denmark; Department of Biomedical Sciences, University of Copenhagen, Copenhagen, Denmark; Centre for Integrated Register-Based Research, CIRRAU, Aarhus University, Aarhus, Denmark; Center for Integrative Sequencing, iSEQ, Aarhus Genome Centre, Aarhus University, Denmark

**Keywords:** Foetal Development, Psychiatry, Bipolar Disorder, Anorexia Nervosa, Mental Health, Developmental Origin of Health and Disease (DOHaD), Gestational Age, Birth Weight, DNA Methylation, Blood, Epigenetic Clock, Biomarker, Twins, Monozygotic, Dizygotic

## Abstract

**Background:** Foetal development indicates the risk of later disease, but has only been associated with few psychiatric disorders. An aggregated molecular marker of development - DNA methylation based estimates of gestational age (DNAmGA) adjusted for GA, can be indicative of foetal health and development. Twins have the same chronological GA and monozygotic (MZ) twins share genetic liability. We leveraged this to examine whether DNAmGA in neonates associate with later psychiatric disorder, independent of chronological GA, maternal characteristics, genetic influences, and shared environmental factors.

**Method:** We estimated DNAmGA in 260 MZ and 396 dizygotic (DZ) twin pairs, later diagnosed with schizophrenia, bipolar disorder, affective/depressive mood disorder, autism spectrum disorder, attention deficit hyperactivity disorder or anorexia. DNAmGA was tested for association with psychiatric outcome by mean discordant twin differences and by linear mixed model (LMM), adjusting for relatedness and potential confounders.

**Results:** We found elevated DNAmGA to associate with anorexia between discordant DZ and MZ twins (0.74 weeks, 95%CI[0.34:1.14] and 0.28 weeks, 95%CI[0.04:0.53], respectively), and with bipolar disorder between discordant MZ twins (0.85 weeks, 95%CI[0.16:1.53]). Elevated DNAmGA associated significantly with both in the LMM analysis (0.56 weeks, 95%CI[0.32:0.83] and 0.89 weeks, 95%CI[0.32:1.51], respectively).

**Conclusions:** Elevated DNAmGA is associated with two later onset psychiatric disorders in twins, and thus supports a developmental origin of disease. This association was not confounded by variation in conventional measures of foetal development nor genetic liability. We therefore propose that a novel molecular marker of development, can differentiate between later psychiatric outcome in newborn twins.

## Background

Gestational development is critical for mental health later in life. In fact, the gestational duration until birth has been associated with mental health, where preterm birth is associated with increased disease risk ^1,2^. Similarly, specific point measures of gestational growth and development, such as birth weight is predictive of both early and later onset mental disorders when normalized for length of gestation^3^ and psychiatric disease in the family (e.g. genetic correlation)^4^.

The latter observation illustrates that proper course of brain development during embryonic life is not only dependent on the length, but also on the quality of gestation, where harmful environmental exposures or genetically programmed disease processes may lead to deviant development^5^. Consistently, gestational exposures to alcohol consumption^6,7^, tobacco smoking ^8^ and stressful events ^9,10^ among others all augment risk of mental disorders. Common genetic factors acting during embryonic brain development further predispose to mental disorders, possibly through reprogramming gene expression patterns^11^ and epigenetics^12^.

This insight has prompted a search for molecular markers of foetal growth and development that reflect effects of both the environmental exposures *in utero* and the genetic background of the individual. Such molecular markers may potentially have the ability to estimate risk of illness and particularly mental disorders, and inform on early pathophysiological processes leading to mental disorders. Recently one such marker, assessing DNA methylation patterns in circulating cells from whole blood, have been shown to correlate with gestational age at birth, termed DNAmGA^13^, analogous to another methylation-based predictor of chronological age in adults^14^. Importantly, the DNAmGA predictor adjusted for gestational age associates with BW and maternal socio-economic status (also a risk factor of adverse mental health outcome in offspring)^13^, as well as maternal depression and child mental health issues^15^ and maternal and child health related factors, such as maternal age and 1-min APGAR score^16^. This suggests that the DNAmGA estimates captures other aspects of developmental processes and neonatal health, in-casu of growth trajectory, in addition to that of gestational length.

In this study, we extend this rational and examined whether DNAmGA estimated at birth can be applied as a marker of specific mental disorders in childhood and adult life. To eliminate uncertainly associated with clinical estimates of gestational age, we perform these analyses in a psychiatric population-based sample of same sex mono- and dizygotic twins, discordant or concordant for six mental disorders as well as in non-affected twin pairs - leveraging the nationwide Danish Neonatal Screening Biobank and detailed diagnostic hospital register-data from the egalitarian and uniform Danish healthcare system.

## Material and Methods

### Sample selection

As a part of the Danish neonatal screening effort for congenital disorders, neonatal heel blood is collected on Guthrie Cards and stored in the Danish Neonatal Screening Biobank (DNSB) at −20°C^17^.

Same sex twin pairs of unknown zygosity were identified by similar criteria of The Lundbeck Foundation Initiative for Integrative Psychiatric Research (iPSYCH) 2012 cohort^18^. In short, selection criteria here were twins chosen among twins born in Denmark May 1, 1981 or later, where both twins in a pair should be alive and resident in Denmark on their 1-year birthday. The twin pairs are born between 1981-2009, where the mother had a Danish social security number, and either one or both of the twins had been diagnosed with schizophrenia (SZ), bipolar disorder (BD), affective/depressive mood disorder (MD) (not including BD), autism spectrum disorder (ASD), attention deficit hyperactivity disorder (ADHD) or anorexia (ANO) by the ICD-10-DCR (F20, F30-F31, F32-F39, F84.0/F84.1/F84.5/F84.8/F84.9, F90.0, F50.0, respectively)^18^ by 2012. A population-based control group of twin pairs, not diagnosed with any of the six disorders (although psychiatric diagnoses may occur) was further included in the dataset. In total 847 same-sex twin pairs were identified. Some individuals had comorbid diagnoses (see Supplementary Figure S1), and in some pairs twins had different diagnoses (e.g. twin/co-twin comorbidities). In both cases samples were used for analysis in more than one psychiatric disorder.

### DNA preparation and microarray

Genomic DNA extraction from neonatal dried blood spots (neoDBS), created from neonatal heel blood soaked Guthrie Cards stored frozen in the DNSB, has previously been described^19^. Briefly, 2×3.2 mm punches of neoDBS were taken for each twin and were used for DNA extraction yielding a sufficient quantity and quality of material (as described by Baekvad-Hansen et al.^20^) for genotyping and DNA methylation profiling^21^. Genomic DNA was extracted with the use of Extract-N-Amp Blood PCR kit (Sigma-Aldrich, St. Louis, Missouri, USA), without further amplification step for Illumina DNAm Array Protocol. Material for genotyping was whole-genome amplified in triplicates with REPLI-g (Qiagen, Hilden, Germany), and analysed according to manufacturer’s instructions using the Infinium PsychChip Array (Illumina, San Diego, California, United States). For DNAm array, genomic DNA was bisulfite-converted using the EZ-96 DNA Methylation Kit (Zymo Research, Irvine, California, USA) according to the manufacturer’s instructions. DNA methylation from the bisulfite-converted DNA was measured using the Infinium HumanMethylation450-BeadChip (450k) or the Infinium HumanMethylationEPIC-BeadChip (EPIC) (see Supplementary Table S1) array (Illumina, San Diego, California, United States) with 7 μL of bisulfite converted genomic DNA, but otherwise according to the manufacturer’s instructions ^22^.

### Data pre-processing and QC

All genetic data was loaded from IDAT intensity files from scanned arrays on iScan (Illumina, San Diego, California, United States). Genotyped SNPs were filtered for MAF>0.05 and CR>0.97. Zygosity of all twin pairs was determined with PLINK v1.07 by Identity by Descent clustering (MZ: PI-hat ≥ 0.9, and DZ: 0.3 ≤ PI-hat ≥ 0.7, for MAF>0.05 variants). This identified 524 same-sex DZ and 323 MZ twin pairs.

Twins were analysed in pairs in two batches (see Supplementary Table S1) on Illumina 450k Array (MZ twins only) or Illumina EPIC Array (MZ, DZ and unaffected twins). Space restriction caused us to not include 53 DZ pairs on DNAm array and these are not used in the subsequent analysis.

DNAm Array IDAT files of 450k and EPIC arrays were loaded separately as red/green intensity datasets (rgSets), with the R-package *Minfi* version 1.24.0 ^23^. 463 DZ and 321 MZ pairs had sufficient quality (Sample call Rate > 0.95 for probe detection P-value < 0.01 and bead count ≥ 3). The rgSets were merged into one combined 450k annotated rgSet, non-shared probes between the two arrays set to missing values and low quality probes (probe call frequency < 0.95, for probe detection P-value < 0.01 and bead count ≥ 3) were also set to missing values.

### Sample filtering

Registry data on GA and BW were obtained from the National Birth Registry, while sampling day was obtained from the DNSB PKU registry. Z-scores of BW for GA were calculated for males and females separately, with the adjustable foetal weight standard for twins published by Zhang et al. ^24^.

Twin pairs were excluded from the study if their blood samples had not been taken on the same day and/or were taken more than 10 days after birth. Samples with insufficient information on BW, GA or lacking verification of individual twin identity within the twin pair were also removed from the study.

After QC and sample filtering 396 same-sex DZ and 260 MZ psychiatric pairs, and 49 same sex DZ and 29 MZ unaffected pairs remained for the analysis (totalling 445 DZ and 289 MZ pairs), as shown in Table 1.

**Table 1:**
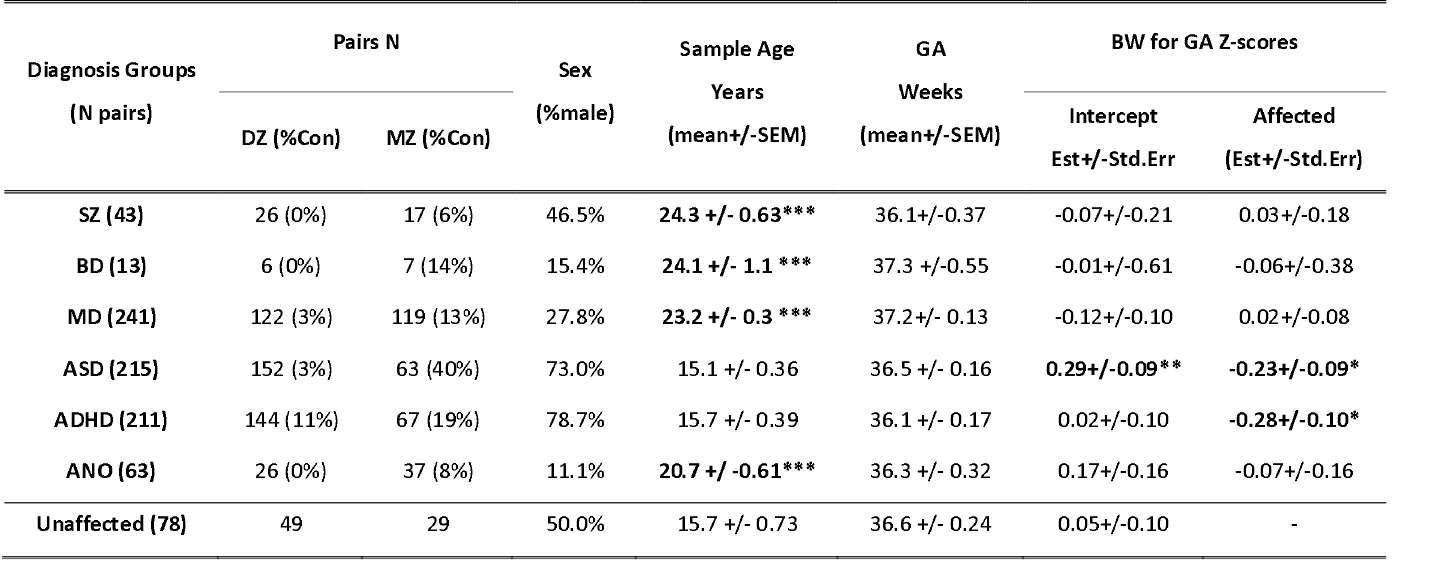
Sample overview for each psychiatric diagnosis group and unaffected twin pairs. Table shows samples size in pairs (N), concordance rate (%Con), sex distribution (%male), mean sample age (at data extraction 2012) and mean gestational age (GA). Differences between each diagnosis group (SZ, BD, MD, ASD, ADHD or ANO) and unaffected pairs were assessed by Welch Two-Sample T-Test (null hypothesis is equal means) for sample age and GA. Samples in diagnosis groups SZ, BD, MD and ANO were older than unaffected when analysed in 2012 (synonymous with earlier birth years), while no significant differences in GA was observed between psychiatric and unaffected samples. BW adjusted for GA (BW for GA) in Z-scores was tested with a LMM controlling for twin pair and zygosity. The “Intercept” column is the estimate of the mean BW for GA of the unaffected co-twins in the affected pairs, while the estimate of the difference in BW for GA of the affected twins from the unaffacted is given in the “Affected” column. Statistical significance is indicated after multiple testing correction with false discovery rate correction (FDR) with FDR alpha: *<0.05, **<0.01 or ***<0.001.

### DNAm-based estimates

#### Knight DNAm GA estimation

DNAmGA estimation was carried out as described by Knight et al. ^13^ using the method available on GitHub (https://github.com/akknight/PredictGestationalAge). Beta values were BMIQ normalized, as optional in the source code, and the 148 CpGs needed for estimation were extracted. 6 of the 148 CpG probes required for the estimator were not indigenous to the EPIC array and thus contained missing values for all samples on both arrays (cg02941816, cg19564877, cg20394284, cg25306927, cg25374854, cg26656135). The analysis was consequently performed on only the 142 CpGs, unless otherwise specified (e.g. in the verification shown in Supplementary Figure S2) as suggested in an application note by the authors (see https://github.com/akknight/PredictGestationalAge). As DNAmGA is largely confounded by GA, we illustrate associations with DNAmGA as DNAmGA Residudals that are linearly adjusted for GA with sample day after birth.

#### Blood Cell Proportion estimates

Blood cell composition (BCC) in the neonatal blood samples were estimated with the Houseman method for DNAm array data, with the neonatal cord blood reference panel from Bakulski et al. ^25,26^. We normalized the reference panel with our DNAm data by the normal-exponential out-of-band (noob) dye bias correction on the rgSet, to correct for background dye bias between datasets (the Bakulski reference panel and our data). Cellular heterogeneity estimates were obtained for B-cells, CD4 T-cells, CD8 T-cells, granulocytes, monocytes, and nucleated red blood cells (Bcells, CD4T, CD8T, Gran, Mono and nRBC, respectively). We did not estimate natural killer cells in the final iteration of our analysis, since in the preliminary analysis this mostly estimated to zero.

### Statistics

All statistical analysis was performed with R Version 3.4.2.

#### Multiple Testing

Tests significance values were corrected for multiple testing bias by applying False Discovery Rate (FDR) correction, and significant p-values were achieved at FDR corrected alpha<0.05.

#### Twin-based heritability and Empirical Bootstrap of Confidence Intervals

We performed 10,000 permutation empirical bootstrapping of the within pair DNAmGA correlation for MZ and DZ twins, and calculated the H^2 by (r(MZ)-r(DZ))*2 for each iteration. 95% CI was determined as the lowest 2.5% value and 97.5% values of the 10,000 permutations.

#### Discordant Twin Model

For each psychiatric diagnosis group (as shown in Table 1) we kept only discordant twin pairs and subtracted DNAmGA of the unaffected twin from the affected twin. The differences were tested with a Student’s T-test of means, with the null hypothesis mean=0, with 95% Confidence interval and p-values.

#### Linear Mixed Model

We applied linear mixed models (LMM) with restricted maximum likelihood (REML), with the R-package lme4 ^27^. This controls for the twin relatedness (pair and zygosity) in the dataset and linearly associates DNAmGA and each psychiatric disorder. We included the twin pair identifier and zygosity as random effects, with intercept varying among zygosity and pair identifier within zygosity as: *(1*|*zygosity/Pair ID)*. We further included the array type (450k/EPIC) as a separate random effect for analysis potentially affected by batch (DNAm array data), accounting for potential the batch effects between the 450k and EPIC array, as: *(1*|*ArrayType)*. In the LMMs we also included potential confounders as covariates – GA, BW for GA, blood cell proportions (common confounder of the heterogeneous blood methylome), birth year, sex and comorbid diagnosis.

#### Permutation Testing for small N

Based on the con/discordant, MZ/DZ twin study design within each diagnosis group we randomly assigned case/control (1/0) status to randomly sampled MZ/DZ psychiatric samples in pairs. We performed 10,000 permutations of each diagnosis group case/control design, as previously described with random effects controlling for relatedness and batch and all covariates. Permuted p-values were obtained as the proportion of permuted p-values smaller than the obtained p-value of the LMM test described above (area under the curve), with the R function *ecdf* (*stats* package).

## Results

### The cohort

We identified 260 monozygotic (MZ) and 396 same-sex dizygotic (DZ) twin pairs that were dis- or concordant for at least one of the six psychiatric diagnoses (SZ, BD, MD, ASD, ADHD, and ANO) by latest 2012, and also included 29 MZ and 49 DZ unaffected twin pairs into our analysis after filtering and quality control (Table 1). A summary of the cohort is given in Table 1, and the occurrence of comorbid diagnoses are illustrated in Supplementary Figure S1. As expected, the twins were born mildly preterm (median of 37 weeks), of which 38% were born prematurely before week 37.

### Size mediates risk of later ASD and ADHD

First, we used a linear mixed model to associate BW-for-GA with each psychiatric disorder. As shown in Table 1, BW-for-GA was significantly lower in the affected compared to the unaffected twins for ASD or ADHD. For ASD it appeared that unaffected co-twins had higher BW-for-GA than the population mean (Z-score of 0), while affected twins were simultaneously smaller.

### DNAm-based estimates of gestational age

Second, we estimated DNAmGA with the method developed by Knight et al. ^13^ and found these to correlate with clinical gestational age plus sample day after birth (r=0.79, N=1468, p-value<2e-16; Supplementary Figure S3a). DNAmGA correlates within twin pairs (r=0.82), and correlation was higher between MZ twins than DZ twins (rMZ=0.88 and rDZ=0.79; Supplementary Figure S3b). Additionally we found DNAmGA adjusted for GA to correlate between both MZ and DZ twins (rMZ=0.65 and rDZ=0.45; Supplementary Figure S3c).

### DNAmGA differences between discordant twins are associated with BD and ANO

Next, we calculated the differences in DNAmGA between discordant twin pairs (ΔDNAmGA = DNAmGA_aff_-DNAmGA_unaff_) and found it to be larger than zero in both MZ and DZ twin pairs discordant for ANO (0.28 weeks, 95%CI [0.04 : 0.53], and 0.74 weeks, 95%CI [0.34 : 1.14], respectively), and in MZ twins pairs discordant for BD (0.85 weeks, 95%CI [0.16 : 1.53]). In combined analysis of MZ and DZ twin pairs, only ANO significantly associated with ΔDNAmGA (0.48 weeks, 95%CI [0.26 : 0.70]). After FDR correction for multiple testing, only ANO remained significantly associated with ΔDNAmGA (P_FDR_= 3.5e-4, see Table 2 and Figure 1, for all analyses). There was no overlap between ANO and BD diagnoses in the two diagnostic groups (Supplementary Figure S1).

**Table 2:** DNAmGA difference of discordant pairs: The table shows the DNAmGA mean difference within discordant pairs for each psychiatric diagnosis (affected – unaffected). Positive values indicate higher DNAmGA in the affected twin relative to the unaffected. This was tested in only MZ pairs, only DZ pairs and in all discordant pairs (MZ and DZ), with One Sample T-Tests of means (null hypothesis is mean=0). Statistical significance was obtained at FDR<0.05. Cursive-bold writing indicates marginal associations that were not significant after FDR correction while bold writing indicates significant association after FDR correction.

| Diagnosis | MZ Discordant Pairs |  |  | DZ Discordant Pairs |  |  | All Discordant Pairs |  |  |
| --- | --- | --- | --- | --- | --- | --- | --- | --- | --- |
|  | N Pairs | Mean [95% CI] | P-Value (FDR) | N Pairs | Mean [95% CI] | P-Value (FDR) | N Pairs | Mean [95% CI] | P-Value (FDR) |
| SZ | 16 | -0.20 [-0.56 : 0.16] | 0.25 (0.51) | 26 | 0.21 [-0.26 : 0.67] | 0.38 (0.56) | 42 | 0.05 [-0.27 : 0.37] | 0.75 (0.75) |
| BD | 6 | 0.85 [0.16 : 1.53] | **0.03 (0.08)** | 6 | 0.20 [-0.91 : 1.30] | 0.67 (0.67) | 12 | 0.52 [-0.05 : 1.09] | 0.07 (0.21) |
| MD | 104 | 0.01 [-0.18 : 0.15] | 0.88 (0.94) | 118 | -0.05 [-0.26 : 0.16] | 0.64 (0.67) | 222 | 0.03 [-0.17 : 0.10] | 0.64 (0.75) |
| ASD | 38 | 0.09 [-0.17 : 0.35] | 0.50 (0.74) | 147 | -0.08 [-0.27 : 0.10] | 0.37 (0.56) | 185 | -0.05 [-0.20 : 0.11] | 0.54 (0.75) |
| ADHD | 54 | 0.01 [-0.27 : 0.25] | 0.94 (0.94) | 128 | 0.12 [-0.08 : 0.33] | 0.24 (0.56) | 182 | 0.08 [-0.08 : 0.24] | 0.31 (0.62) |
| ANO | 34 | 0.28 [0.04 : 0.53] | **0.03 (0.08)** | 26 | 0.74 [0.34 : 1.14] | **7.7e-4 (4.6e-3)** | 60 | 0.48 [0.26 : 0.70] | **5.8e-5 (3.5e-4)** |

**Figure 1:**
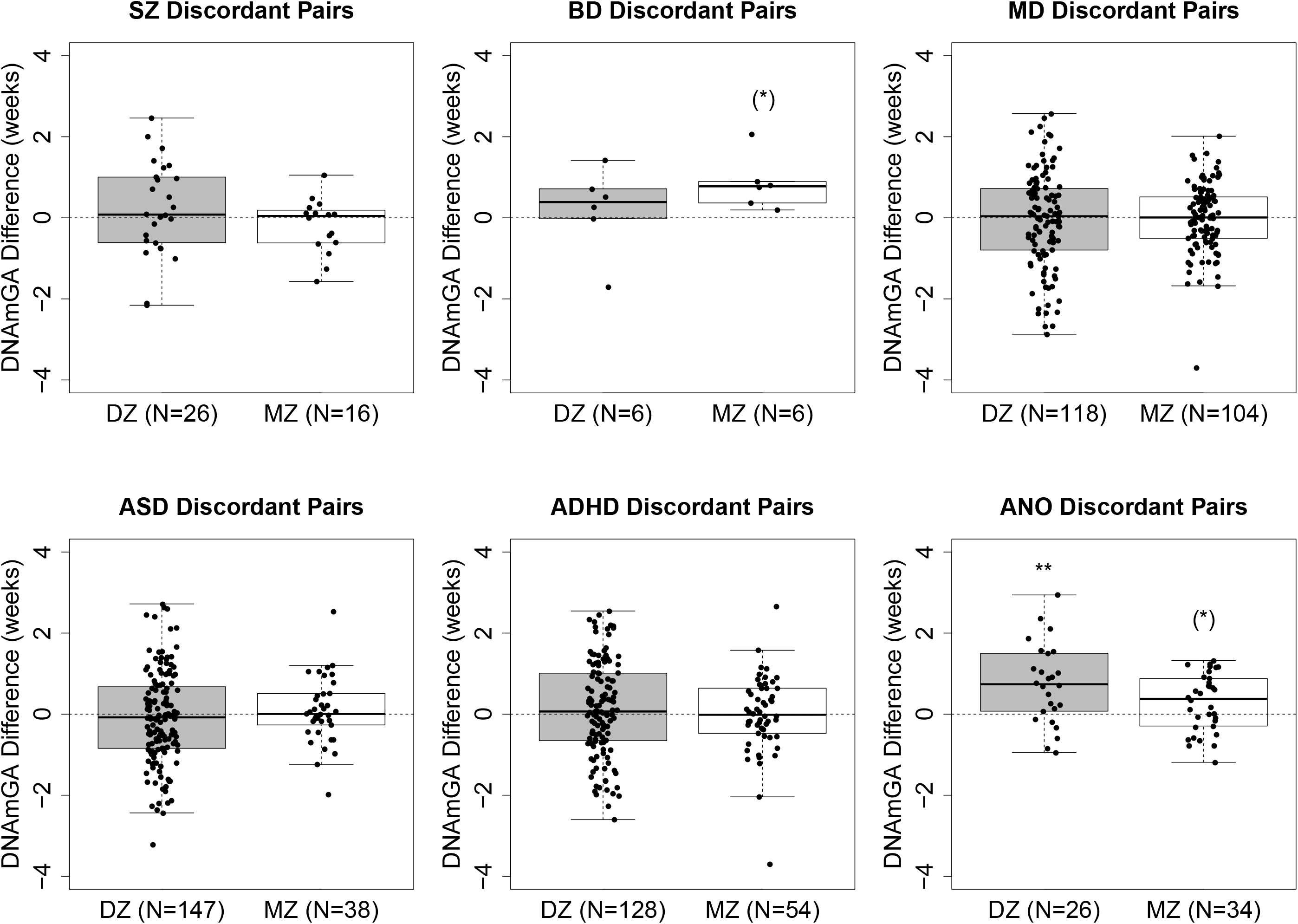
Boxplots of DNAmGA difference in psychiatric discordant pairs. Boxplot shows distribution of DNAmGA differences (affected – unaffected) within psychiatric discordant pairs for each psychiatric diagnosis. Differences within DZ pairs are illustrated with gray boxes while differences within MZ pairs are illustrated with white boxes. The sample size is given for each box as N pairs and difference between each twin pair is illustrated with black jittered dots. Dashed line marks a difference of 0, and dots above the line are interpreted as elevated DNAmGA in the affected twin over the unaffected co-twin. Test P-values from One-Sample T-Test (null hypothesis is mean=0) are illustrated as (*)<0.1, *<0.05, **<0.01, after FDR correction for multiple testing.

### Elevated DNAmGA is associated with BD and ANO

Finally, to allow for adjustment for potential within- and between-pair confounders, including relatedness, we used a linear mixed model (LMM) approach to analyse DNAmGA in all MZ and DZ pairs, con- and discordant of psychiatric diagnosis in each diagnostic group, as well as unaffected pairs (see Table 3). In this model we adjusted DNAmGA for GA, sample day, BW-for-GA, DNAm based blood cell composition, comorbid diagnoses and twin comorbidity, and for birth year and sex. As shown in Table 3, elevated DNAmGA associated with ANO (0.56 weeks, 95%CI [0.32:0.83], P_FDR_ = 0.00014) and with BD (0.89 weeks, 95%CI [0.32:1.51], P_FDR_ = 0.012) (Figure 2), while it did not associate with any other diagnosis (Table 3). We note that, although BW-for-GA, sex and birth year associate with DNAmGA adjusted for GA (Supplementary Figures S4-6), only adjusting for GA and sample day provided comparable test results (The P-value of this is illustrated in Supplementary Figure S7). To test the accuracy of the asymptotic p-values from the LMM, we performed permutation tests for each diagnostic group, confirming the validity for small sample sizes and case-control ratio (Supplementary Figure S7 and Table 3).

**Table 3:** Summarized results from LMMs of DNAmGA association with psychiatric diagnosis adjusted for Comorbid diagnosis, GA, sample day, sex, birth year, BW for GA and blood cell composition). Table summarizes the number of affected and unaffected samples (N) analysed in each model. Unaffected samples included 156 unaffected samples from the unaffected pairs. Comorbid diagnosis of affected twin or of the co-twin were included as covariates in the LMM and are listed for each model. Covariates included in all LMMs were BW for GA, blood cell composition, GA, sample day, birth year and sex, and we adjusted for twin pair, zygosity and array type with random effects. LMM estimates with 95% CI of DNAmGA effect size in weeks, P-value (with FDR correction for multiple testing and permuted p-values) are listed for each diagnosis. Statistical significance was obtained at FDR<0.05. Bold writing indicates significant association after FDR correction. Significance was confirmed by permuted p-values.

| Diagnosis Group | Affected (N samples) | Unaffected (N samples) | Comorbid Diagnoses (in twin/co-twin) | Est (weeks) [95%CI] | LMM p-value / (FDR) / [PermP.] |
| --- | --- | --- | --- | --- | --- |
| SZ | 44 | 198 | BD, MD, ASD, ADHD, ANO | -0.13 [-0.52:0.27] | 0.52 (0.77) [0.53] |
| BD | 14 | 168 | SZ, ADHD | 0.89 [0.32:1.51] | **0.004 (0.012) [0.0035]** |
| MD | 260 | 378 | SZ, ASD, ADHD, ANO | 0.02 [-0.12:0.16] | 0.81 (0.81) [0.81] |
| ASD | 245 | 341 | SZ, MD, ADHD, ANO | -0.02 [-0.17:0.13] | 0.76 (0.81) [0.77] |
| ADHD | 240 | 338 | SZ, BD, MD, ASD | 0.09 [-0.07:0.25] | 0.27 (0.54) [0.26] |
| ANO | 66 | 216 | SZ, MD, ASD | 0.56 [0.32:0.83] | **0.000024 (0.00014) [<1e-4]** |

**Figure 2:**
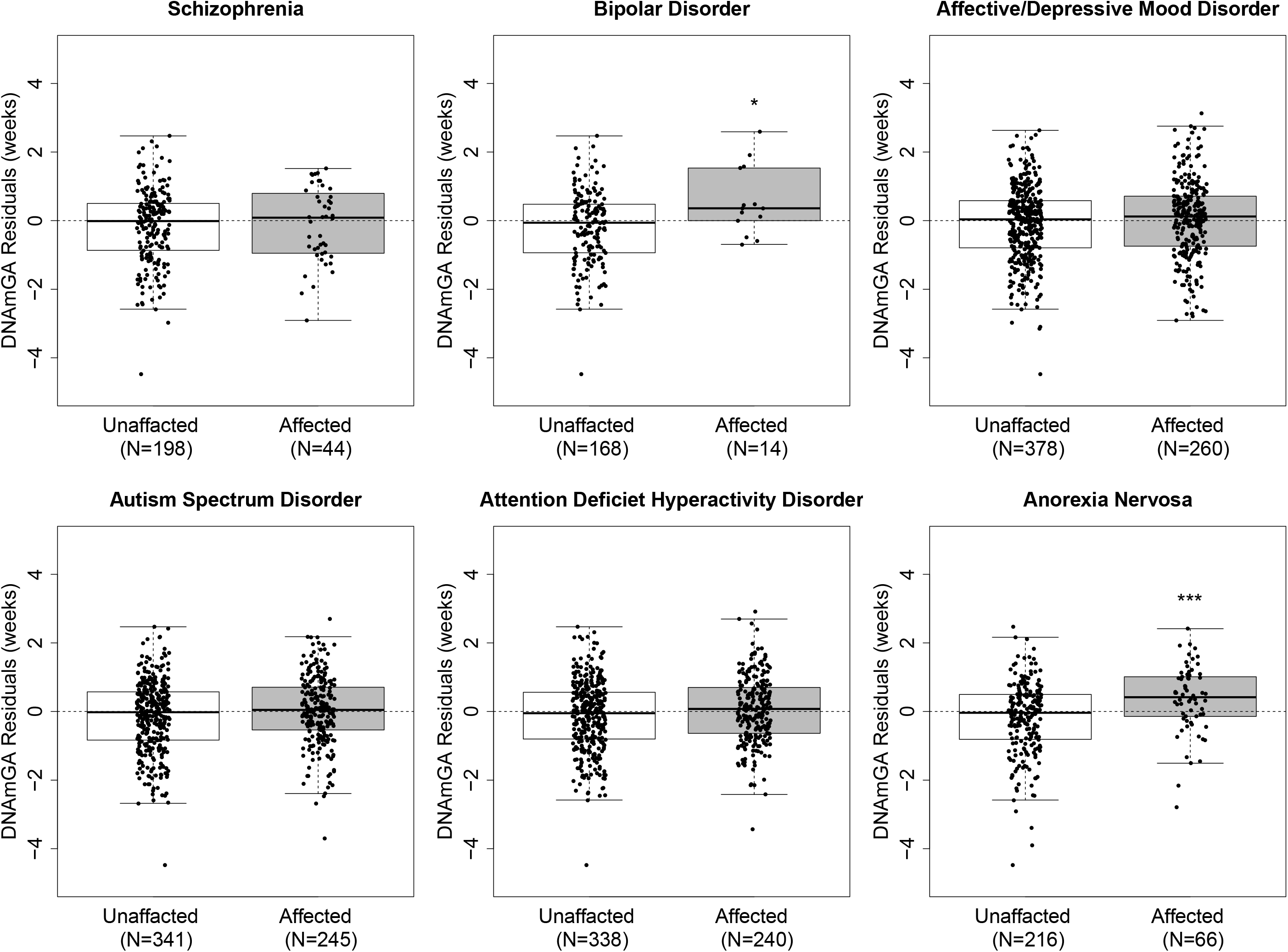
Increased DNAmGA adjusted for within and between pair covariates (shown as residuals) associates with later BD and ANO. Boxplots shows DNAmGA adjusted for GA, sample day, sex, birth year, BW for GA and blood cell composition as residuals association with each psychiatric disorder. Unaffected (white box) and affected (gray box) samples of each disorder are illustrated as jittered dots. Dashed line marks DNAmGA residuals=0. LMM FDR corrected p-values are illustrated as *<0.05, **<0.01, ***<0.001.

## Discussion

Here we show that a molecular marker of foetal development (DNAmGA) is associated with at least two later onset psychiatric disorders. This is, to our knowledge, the first demonstration of a developmental origin of BD and ANO, that is independent of gestational duration.

We used a blood DNA-methylation-based estimator of GA that also correlates with other birth measures of development and health, e.g. BW and APGAR. While these measures correlate with GA, they are not fully explained by it (e.g. variance in BWforGA), and thus may represent alterations in developmental trajectories that are associated with foetal health. We therefore suggest that DNAmGA adjusted for GA can be interpreted as an accumulative score of multiple physiological processes affecting neonatal health and development. We suggest that DNAmGA can be used as a molecular marker of development, without the shortcomings of conventional surrogate markers, although further studies in large and independent samples are necessary to support this hypothesis.

Our findings suggest that early and *in utero* exposures influencing (brain) development may act through the epigenome and regulate gene expression^12^, which is also supported by our earlier findings in neonatal blood showing that differential DNAm in genes involved with foetal brain development and neurogenesis is associated with later psychiatric outcome in individuals with 22q11.2 deletion syndrome^28^.

The twin design applied in this study has a number of important advantages as chronological GA is identical for a pair of MZ twins, which eliminates the uncertainty of conventional methods to estimate and correct for clinical GA estimates that rely on e.g. last menstrual period or ultrasound assessed foetal metrics. Moreover, clinical GA estimates are influenced by several maternal characteristics, (i.e. smoking and obesity^29^) and by genetic factors^30^. Thus differences in DNAmGA in a twin pair will primarily reflect non-shared environment and stochastic effects for MZ twins, but also has a genetic components in DZ twins – as demonstrated in this study by twin heritability estimates of DNAmGA.

We confirmed the results obtained from the simple discordant-twin model in MZ twins, using a Linear Mixed Model that allowed us to adjust for potential confounders, e.g. blood cell composition. This is relevant because it indicates that the methylation differences do not reflect a global confounding by variance in blood cell type heterogeneity.

Another study of DNAmGA in children of mothers with antenatal depression demonstrated that decreased DNAmGA was associated with later psychiatric problems^31^. While such studies in singletons are prone to be influenced by underlying genetic and maternal characteristics influencing estimate, as well as uncertainty in measures of chronological GA, we do not expect them to reflect the same pathophysiology as observed in this study of twins.

Our study is generally limited by the small sample sizes which hamper the power to detect other possible associations than those for BD and ANO. This emphasizes the need for replication in larger independent cohorts which would benefit from the inclusion of both twins and singletons. The latter acknowledging that twins may not represent the pathophysiology of singletons as they are associated with multiple birth complications, prematurity and lower BW.

In conclusion, we show for the first time, that an epigenetic measure of development measured at birth, DNAmGA, is indicative of risk for developing BD and ANO in adulthood. This risk is independent of genetic influences and conventional measures of neonatal development (i.e., GA, BW for GA). Thus, our findings further support a developmental origin of psychiatric disorders.

## Data Availability

Relevant summary stats and other aggregated values can be freely shared.
Access to raw data is possible but due to the sensitive nature of the data only in secured environments established by the authors.
contract the corresponding with access requests.

## Declarations

### Ethics approval and consent to participate

The Danish Scientific Ethics Committee (approval number 37356) approved this study, as did the Danish Health Data Authority, the Danish Data Protection Agency and the Danish Neonatal Screening Biobank steering committee. Consent is given in the form of passive consent, and relies on protection of personal information and anonymization of all involved personal data and sample material.

The manuscript was made available on bioRxiv ahead of publishing.

### Consent for publication

Not applicable

### Availability of data and material

Individual level data can be accessed only through secure servers operated by iPSYCH consortium. Download of individual level information is prohibited due to the sensitive nature of these data. iPSYCH encourage national and international collaboration. For details, please contact the corresponding authors.

## Authors’ contributions

CSH carried out all QC and data analysis of methylation, interpretation of results and prepared the manuscript. AS, AWD and SW provided interpretation of methylation analysis in addition AWD provided pre-processing pipeline and AS provided epigenetic psychiatric interpretation. DH provided vital insight into twin analysis and interpretation of results and SH provided interpretation of these from DNA methylation perspective. MABH, DMH and JBG arrayed and selected neoDBS samples from the DNSB (with our lab technician team - see acknowledgements) and provided vital background knowledge and protocol for the work with such material. MC provided vital knowledge of pregnancy, delivery and neonatal health and development in Denmark reflected in the DNSB and provided vital interpretation of the results in this paper. PBM, CBP and MGP provided and curated all psychiatric phenotypes and registry data in the cohort and provided all additional twin relevant information for verification of the pairs. CL provided vital intellectual input on project feasibility, design and the application of DNA methylation in multifactorial disorders. TMW, JBG and AB conceived and designed the study as well as interpreted results and formed conclusions. Specifically AB provided the concept, analysis pipeline and conceived additional supportive analysis in heritability estimates, linear mixed model and interpretation.

All authors read and approved the final manuscript.

## Acknowledgements

This research has been conducted using the Danish National Biobank resource, supported by the Novo Nordisk Foundation.

Our talented team of technicians: Marie Louise Johannsen, Rikke Holm Agerbo, Mia Egeberg Engwald and Gimel Pucci Infante Jørgensen, without whom this study would not have been feasible in such a large complex cohort, where high data quality from small input amounts is a necessity, and the many twin pairs need to be correctly identified and carefully extracted from the DNSB.

This publication would not have been possible without the passionate support and ideas of our dear departed friend and colleague Mads Vilhelm Hollegaard. He envisioned and paved the way for this and many other projects rooted in the DNSB – he is missed immensely.

